# The Effects of a Digital Exercise Therapy Application for Patients with Shoulder and Knee Pain During the COVID-19 Pandemic

**DOI:** 10.64898/2026.09.25.26364013

**Authors:** Kevin H. McLaughlin, Marc Gruner, Negar Ahmadian, Tiffany Patch, Bashir Zikria, Richard L. Skolasky

## Abstract

**Objective:** To examine changes in function and pain interference among patients utilizing a digital exercise therapy application (DETA) for knee and shoulder pain.

**Methods:** We conducted a retrospective analysis of data from patients utilizing a DETA for non-operative knee and shoulder pain. Primary outcomes for the study were 8-week changes in physical function, measured using the PROMIS-Physical Function (knee pain) and PROMIS-Upper Extremity Function (shoulder pain); and pain interference, measured using the PROMIS-Pain Interference. Data on patient characteristics was used to examine outcomes among specific patient subgroups.

**Results:** We observed a 3.5 point (SD: 7.8) increase in function and a 3.9 point (SD: 7.5) decrease in pain interference overall (p<0.001), with changes in function and pain interference surpassing the minimal clinically important difference for patients with knee and shoulder pain. Patients with knee pain experienced greater improvements in function than those with shoulder pain (p=0.02), as did patients who report moderate-high levels of exertion during exercise compared to those reporting low levels of exertion (p=0.05). We saw greater reductions in pain interference among those with higher education (p=0.03), those with more acute symptoms (p=0.01) and those reporting moderate-high levels of exercise exertion (p=0.02).

**Conclusion:** We observed a clinically meaningful improvement in pain interference and function among patients utilizing a DETA for knee and shoulder pain. Patients with knee pain, higher levels of education, more acute symptoms, and those who reported higher levels of exertion with exercise experienced the greatest changes in function and pain.

**Impact Statement:** This study indicates that self-guided interventions, such as those delivered through a DETA, may be beneficial for patients with knee and shoulder pain. These results may also inform the development of patient profiles that can be used to identify patients most appropriate to receive care through a mobile application.

## Introduction

Prior to the COVID-19 pandemic, telerehabilitation delivered by physical therapists was sparsely utilized due to a number of barriers surrounding policy and reimbursement.^1^ However, during the pandemic, the need to deliver physical therapy care while reducing direct contact between patients and providers led to changes in policy and reimbursement that facilitated the rapid expansion of telerehabilitation. Since that time, telerehabilitation has been shown to be acceptable among patients and providers, with early evidence suggesting that telerehabilitation may provide similar outcomes to in-person physical therapy for patients with musculoskeletal pain.^2-4^

During its initial expansion in early 2020, telerehabilitation largely referred to physical therapy visits completed via real-time video visits. Since that time, emerging technology and evolutions in healthcare policy have facilitated the continued expansion of telerehabilitation, which now includes improved methods of patient-provider communication between visits, the ability of physical therapists to monitor patient outcomes remotely, and to deliver interventions (e.g., exercise) asynchronously through mobile applications. In particular, the use of mobile applications to deliver asynchronous care stands to greatly improve access to physical therapy, reduce the cost of care, and expand the bandwidth of individual physical therapists.

Delivering asynchronous care using a mobile application allows patients to perform a semi-independent rehabilitation program at a time and in a place that is most convenient for them, without the challenges associated with scheduling in-clinic of telehealth appointments. Early evidence also suggests that care delivered through a mobile application is effective in reducing pain and disability among patients with musculoskeletal pain, including those with back, neck, knee, shoulder, and hip pain.^5-9^ However, evidence in this area is limited and more research is needed to further examine the effectiveness of this approach. In addition, it is likely that certain patients will respond more readily to care provided through a mobile application, an area that needs further exploration before this approach can be fully integrated into physical therapy care models. As such, the purpose of this study was to examine the effects of a digital exercise therapy application (DETA) delivered using a mobile application for patients with knee and shoulder pain presenting to a sports medicine clinic during the early phases of the COVID-19 pandemic. In addition, we sought to examine treatment effects among specific patient subgroups to begin identifying the types of patients who might respond best to this treatment approach.

## Methods

This study is a retrospective analysis of longitudinal data collected among patients utilizing a DETA as part of their routine non-operative clinical treatment for knee and shoulder pain during the COVID-19 pandemic.

### Patients

When outpatient physical therapy visits were limited at our institution to reduce COVID-19 transmission, sports medicine physicians and orthopaedic surgeons at our institution began to offer access to a DETA for patients who would traditionally be referred to physical therapy. Since then, they have continued to offer this alternative to patients who are unable to or prefer not to attend physical therapy but may benefit from therapeutic exercise. This study includes adults (≥ 18) electing to utilize a DETA for knee or shoulder pain between May 2020 and May 2022. Our cohort did not include individuals receiving surgery as these patients are typically seen in-person for post-operative rehabilitation.

### Interventions

All patients in this study utilized a DETA delivered through a mobile application (Limber Health”) specifically designed for patients with musculoskeletal pain. Upon enrollment on the DETA platform, patients completed a series of questionnaires assessing baseline physical and psychosocial factors, which were then used to match patients with an initial exercise program. These exercises were then progressed based on feedback provided by the patient within the application regarding difficulty with exercises, pain experienced during exercise, and number of exercise sessions completed. All exercises were accompanied with pictures, videos, and written instructions. The DETA used in this study has been described previously.^6^

### Outcomes

Our primary outcomes in this study were 8-week changes in physical function and pain interference as measured by the Patient-Reported Outcomes Measurement Information System (PROMIS). For patients with knee pain, function was measured using the PROMIS-Physical Function (PROMIS-PF) V2.0. The PROMIS-PF is a computer adaptive test (CAT) that measures patient-reported function, which are reported on a T-scale (0-100), with higher scores representing greater function and a score of 50 representing means levels of function among the general adult population.^10^ The minimal clinically important difference (MCID) on the PROMIS-PF for patients with knee pain has been reported to be 2.2 points.^11^ For patients with shoulder pain, function was measured using the PROMIS-Upper Extremity Function (PROMIS-UE) which is similar to the PROMIS-PF but is comprised of items specifically designed to assess upper extremity function.^12^The PROMIS-UE has an MCID of 2.1 points for patients with upper extremity pain.^13^ Pain interference was measured for all patients using the PROMIS-Pain Interference (PROMIS-PI) where higher scores indicated greater levels of pain interference and a score of 50 represents mean levels of pain interference among a population consisting of healthy individuals and those with chronic medical conditions.^14^ The PROMIS-PI has an MCID of 2.0 points for those with musculoskeletal pain.’^11^ All PROMIS CAT measures have a standard deviation of 10 points.

We included several patient characteristics in our analysis to examine whether certain types of patients responded differently to the DETA intervention than others. Our dataset included patient demographics, as well as the length of time their condition had been affecting them, how frequently patients exercised and their level of exertion during exercise.

### Data Analysis

Changes in function and pain interference were calculated based on the difference between baseline and 8-week PROMIS measures, with paired t-tests used to examine differences between baseline and 8-week scores. To explore potential relationships between patient characteristics and response to DETA intervention, we report changes in function and pain interference stratified by patient characteristics, including age, body region, BMI, education, chronicity and exercise habits. Differences in 8-week changes between patient characteristic groups were examined using t-tests for 2 group comparisons and ANOVA for ≥ 3 group comparisons. Differences with a p-value of less than 0.05 were considered significant.

### Ethics Approval

This study was approved by the Johns Hopkins Medicine Institutional Review Board.

## Results

We identified 231 patients for inclusion in our cohort, including 120 (52%) patients with knee pain and 111 patients (48%) patients with shoulder pain **(Table 1)**. The average age of those in our cohort was 53.7 (SD: 16.1) years, slightly under half (44%) were female, with the majority of patients self-identifying as White (68%) and non-Hispanic (90%).

**Table 1.**
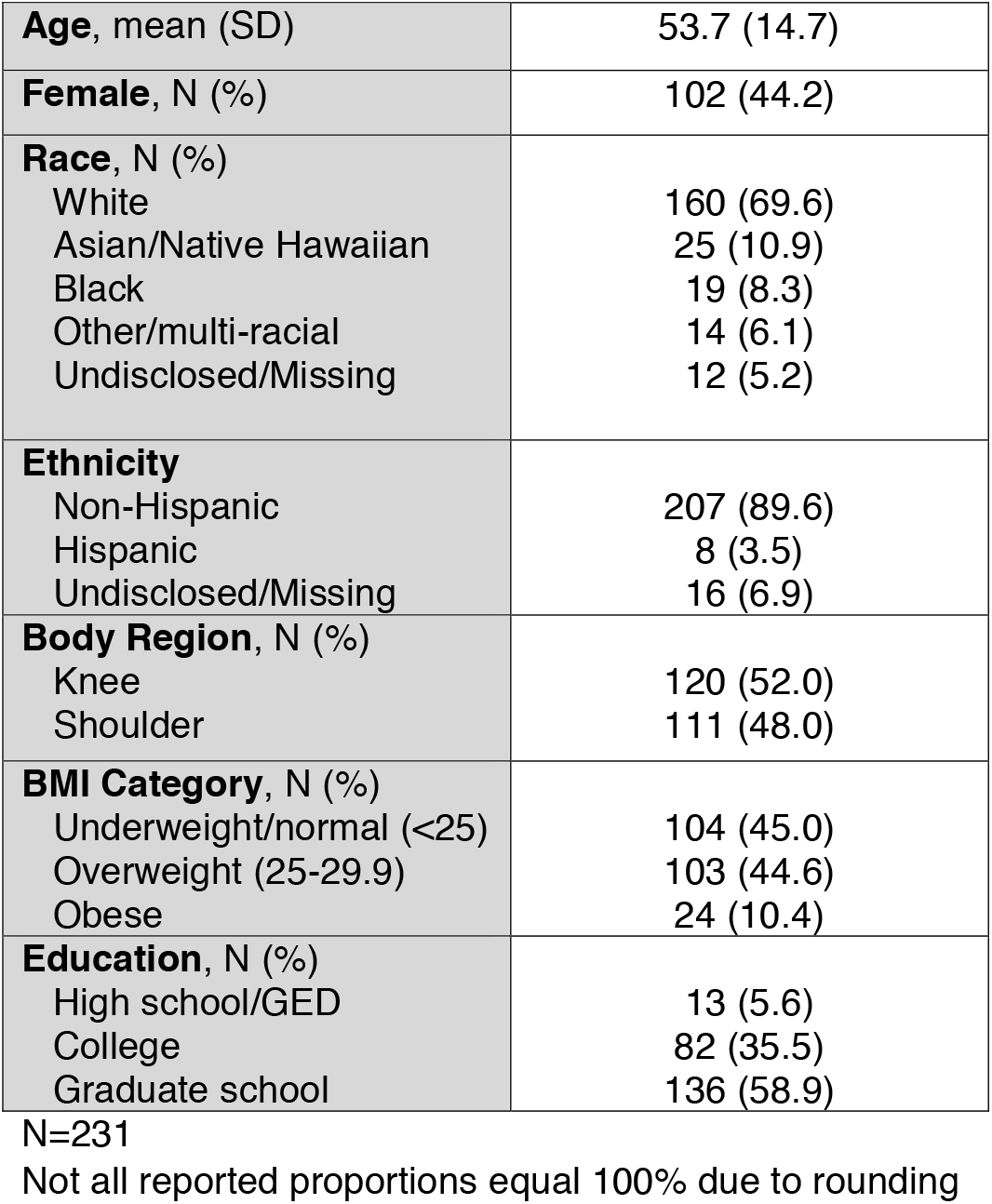
Patient Demographics.

|  |  |
| --- | --- |
| **Age**, mean (SD) | 53.7 (14.7) |
| **Female**, N (%) | 102 (44.2) |
| **Race**, N (%) |  |
| White | 160 (69.6) |
| Asian/Native Hawaiian | 25 (10.9) |
| Black | 19 (8.3) |
| Other/multi-racial | 14 (6.1) |
| Undisclosed/Missing | 12 (5.2) |
| **Ethnicity** |  |
| Non-Hispanic | 207 (89.6) |
| Hispanic | 8 (3.5) |
| Undisclosed/Missing | 16 (6.9) |
| **Body Region**, N (%) |  |
| Knee | 120 (52.0) |
| Shoulder | 111 (48.0) |
| **BMI Category**, N (%) |  |
| Underweight/normal (<25) | 104 (45.0) |
| Overweight (25-29.9) | 103 (44.6) |
| Obese | 24 (10.4) |
| **Education**, N (%) |  |
| High school/GED | 13 (5.6) |
| College | 82 (35.5) |
| Graduate school | 136 (58.9) |

N=231 Not all reported proportions equal 100% due to rounding

We observed significant differences in pain interference and function between baseline and 8-week follow up for patients with shoulder pain and knee pain **(Figure 1)**. The mean decrease in pain interference was 3.1 points (p <0.001) for patients with shoulder pain and 4.6 points (p <0.001) for patients with knee pain. The mean increase in physical function was 2.2 points (p=0.003) for patients with shoulder pain and 4.7 points (p< 0.001) for those with knee pain. Changes in pain interference and function surpassed published values for MCID in both regions.

**Figure 1.**
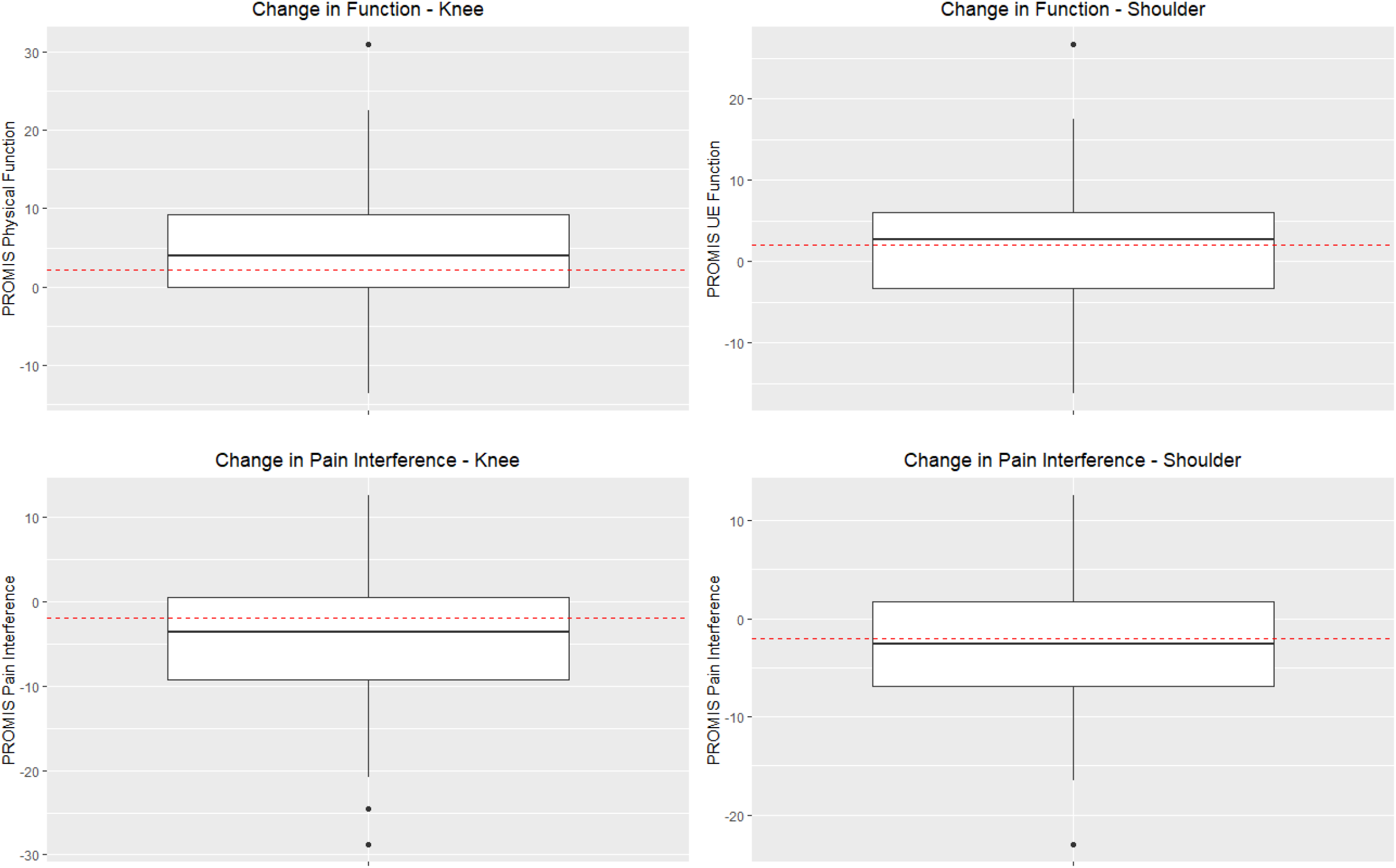
Eight-Week Changes in PROMIS Measures by Body Region. Red dashed line represents the minimal clinically important difference for the PROMIS measure and body region displayed in each graph

We identified significant differences in 8-week outcomes between multiple patient characteristic groups **(Table 2)**. For example, we identified that patients with knee pain reported significantly greater improvements in function (4.7 points [SD: 7.7]) compared to those with shoulder pain (2.2 points [SD: 7.8]) (p=0.02). We also observed that patients with college or graduate school education experiences greater reductions in pain interference than those who reported high school/GED-level education (p=0.03). Patients with acute conditions (<1 month) reported greater reductions in pain interference than those with more chronic symptoms (p=0.01). Lastly, patients who reported moderate and high levels of exertion during exercise reported greater reductions in pain interference (p=0.02) and greater improvements in function (p=0.05) than those who reported low levels of exercise exertion.

**Table 2.**
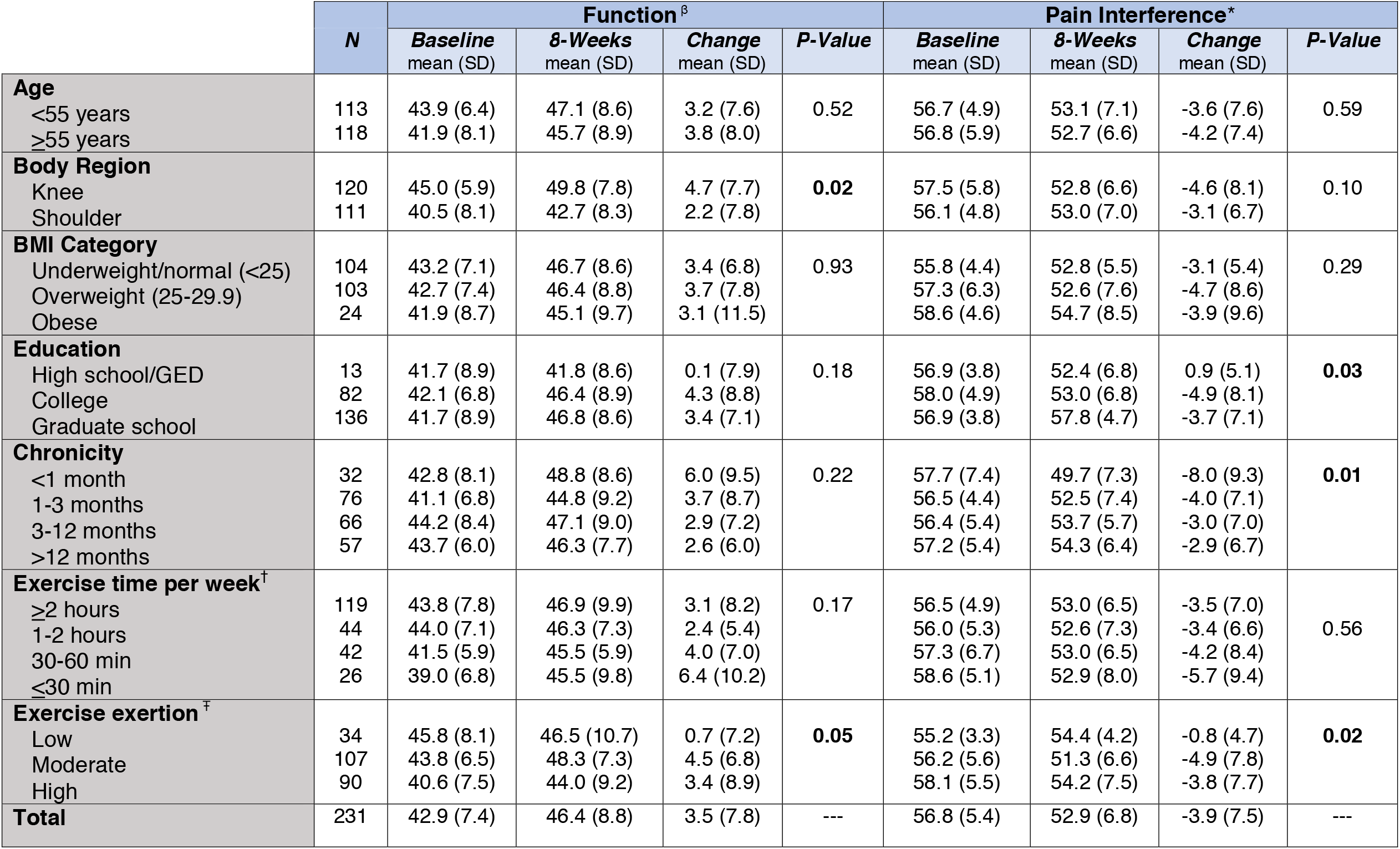
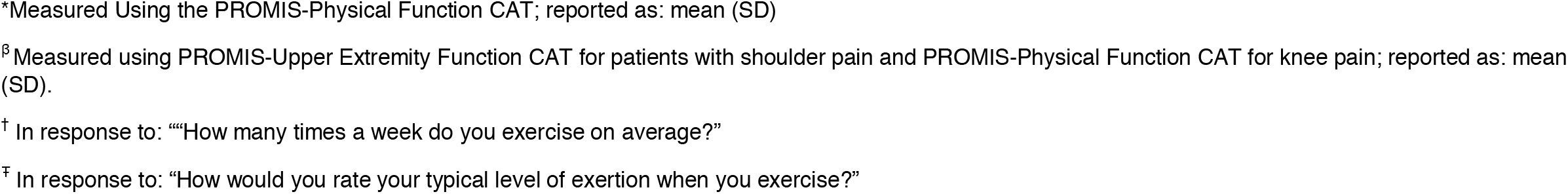
Relationship Between Baseline Factors and Changes in Pain and Function.

|  | *N* | Function<sup>β</sup> |  |  |  | Pain Interference* |  |  |  |
| --- | --- | --- | --- | --- | --- | --- | --- | --- | --- |
|  |  | Baseline<br>mean (SD) | 8-Weeks<br>mean (SD) | Change<br>mean (SD) | *P-Value* | Baseline<br>mean (SD) | 8-Weeks<br>mean (SD) | Change<br>mean (SD) | *P-Value* |
| **Age** |  |  |  |  |  |  |  |  |  |
| <55 years | 113 | 43.9 (6.4) | 47.1 (8.6) | 3.2 (7.6) | 0.52 | 56.7 (4.9) | 53.1 (7.1) | -3.6 (7.6) | 0.59 |
| ≥55 years | 118 | 41.9 (8.1) | 45.7 (8.9) | 3.8 (8.0) |  | 56.8 (5.9) | 52.7 (6.6) | -4.2 (7.4) |  |
| **Body Region** |  |  |  |  |  |  |  |  |  |
| Knee | 120 | 45.0 (5.9) | 49.8 (7.8) | 4.7 (7.7) | **0.02** | 57.5 (5.8) | 52.8 (6.6) | -4.6 (8.1) | 0.10 |
| Shoulder | 111 | 40.5 (8.1) | 42.7 (8.3) | 2.2 (7.8) |  | 56.1 (4.8) | 53.0 (7.0) | -3.1 (6.7) |  |
| **BMI Category** |  |  |  |  |  |  |  |  |  |
| Underweight/normal (<25) | 104 | 43.2 (7.1) | 46.7 (8.6) | 3.4 (6.8) | 0.93 | 55.8 (4.4) | 52.8 (5.5) | -3.1 (5.4) | 0.29 |
| Overweight (25-29.9) | 103 | 42.7 (7.4) | 46.4 (8.8) | 3.7 (7.8) |  | 57.3 (6.3) | 52.6 (7.6) | -4.7 (8.6) |  |
| Obese | 24 | 41.9 (8.7) | 45.1 (9.7) | 3.1 (11.5) |  | 58.6 (4.6) | 54.7 (8.5) | -3.9 (9.6) |  |
| **Education** |  |  |  |  |  |  |  |  |  |
| High school/GED | 13 | 41.7 (8.9) | 41.8 (8.6) | 0.1 (7.9) | 0.18 | 56.9 (3.8) | 52.4 (6.8) | 0.9 (5.1) | **0.03** |
| College | 82 | 42.1 (6.8) | 46.4 (8.9) | 4.3 (8.8) |  | 58.0 (4.9) | 53.0 (6.8) | -4.9 (8.1) |  |
| Graduate school | 136 | 41.7 (8.9) | 46.8 (8.6) | 3.4 (7.1) |  | 56.9 (3.8) | 57.8 (4.7) | -3.7 (7.1) |  |
| **Chronicity** |  |  |  |  |  |  |  |  |  |
| <1 month | 32 | 42.8 (8.1) | 48.8 (8.6) | 6.0 (9.5) | 0.22 | 57.7 (7.4) | 49.7 (7.3) | -8.0 (9.3) | **0.01** |
| 1-3 months | 76 | 41.1 (6.8) | 44.8 (9.2) | 3.7 (8.7) |  | 56.5 (4.4) | 52.5 (7.4) | -4.0 (7.1) |  |
| 3-12 months | 66 | 44.2 (8.4) | 47.1 (9.0) | 2.9 (7.2) |  | 56.4 (5.4) | 53.7 (5.7) | -3.0 (7.0) |  |
| >12 months | 57 | 43.7 (6.0) | 46.3 (7.7) | 2.6 (6.0) |  | 57.2 (5.4) | 54.3 (6.4) | -2.9 (6.7) |  |
| **Exercise time per week<sup>†</sup>** |  |  |  |  |  |  |  |  |  |
| ≥2 hours | 119 | 43.8 (7.8) | 46.9 (9.9) | 3.1 (8.2) | 0.17 | 56.5 (4.9) | 53.0 (6.5) | -3.5 (7.0) | 0.56 |
| 1-2 hours | 44 | 44.0 (7.1) | 46.3 (7.3) | 2.4 (5.4) |  | 56.0 (5.3) | 52.6 (7.3) | -3.4 (6.6) |  |
| 30-60 min | 42 | 41.5 (5.9) | 45.5 (5.9) | 4.0 (7.0) |  | 57.3 (6.7) | 53.0 (6.5) | -4.2 (8.4) |  |
| ≤30 min | 26 | 39.0 (6.8) | 45.5 (9.8) | 6.4 (10.2) |  | 58.6 (5.1) | 52.9 (8.0) | -5.7 (9.4) |  |
| **Exercise exertion<sup>‡</sup>** |  |  |  |  |  |  |  |  |  |
| Low | 34 | 45.8 (8.1) | 46.5 (10.7) | 0.7 (7.2) | **0.05** | 55.2 (3.3) | 54.4 (4.2) | -0.8 (4.7) | **0.02** |
| Moderate | 107 | 43.8 (6.5) | 48.3 (7.3) | 4.5 (6.8) |  | 56.2 (5.6) | 51.3 (6.6) | -4.9 (7.8) |  |
| High | 90 | 40.6 (7.5) | 44.0 (9.2) | 3.4 (8.9) |  | 58.1 (5.5) | 54.2 (7.5) | -3.8 (7.7) |  |
| **Total** | 231 | 42.9 (7.4) | 46.4 (8.8) | 3.5 (7.8) | --- | 56.8 (5.4) | 52.9 (6.8) | -3.9 (7.5) | --- |

\*Measured Using the PROMIS-Physical Function CAT; reported as: mean (SD)
<sup>β</sup> Measured using PROMIS-Upper Extremity Function CAT for patients with shoulder pain and PROMIS-Physical Function CAT for knee pain; reported as: mean (SD).
<sup>†</sup> In response to: “How many times a week do you exercise on average?”
<sup>‡</sup> In response to: “How would you rate your typical level of exertion when you exercise?”

## Discussion

We observed that patients utilizing a DETA experienced significant improvement in pain interference and physical function among patients with non-operative knee and shoulder pain. Importantly, the average improvements in physical function and pain interference reported by patients in this study surpassed published MCID values, indicating clinically meaningful improvements. Additionally, our results indicate that certain types of patients, such as those with higher levels of education, acute symptoms and those who exercise with higher levels of exertion responded more positively to a DETA than others. We also observed that changes in function and pain interference were larger among those with knee pain compared to those with shoulder pain. The results of this study have important implications for the use of DETAs for patients with knee and shoulder pain and may inform the development of future studies in this area, which are needed.

The results of this study align with those of previous studies that have examined the use of a DETA for patients with knee pain. For example, a randomized clinical trial comparing the effects of DETA to conventional physical therapy (N=60), found that patients in the DETA group experienced significantly greater improvements in physical function and reductions in pain interference compared to the conventional physical therapy group.^6^ The authors of the previous study reported greater improvements in physical function (6.0 ± 6.6 versus 4.5 ± 7.7) and pain interference (−6.1 ± 6.7 versus -4.5 ± 8.0) among patients in their study compared to those with knee pain in our study. It is possible that these changes are related to the acuity of patients’ symptoms in the previous study, which was not reported, or the level of patient engagement (e.g., number of exercise sessions completed) with the DETA which was high in the previous study but not included in the current study. However, these studies taken together indicate that a DETA may be an effective approach for patients with knee pain. Larger scale studies are needed to adequately assess the utility of this treatment approach for this patient population.

The effects of digital care programs have also been examined for those with shoulder pain. Janela and colleagues^7^ conducted a single-arm interventional cohort study to examine the use of a digital care program for those with chronic shoulder pain. The authors of that study reported an average reduction in disability of 51.6% (13.45 points) over a 12-week intervention period, measured by the Quick Disabilities of the Arm, Shoulder and Hand (QuickDASH) questionnaire. The use of different functional outcomes between the previous study and the current study limits our ability to directly compare outcomes reported by each study. However, it is likely that that the changes reported in the previous study are larger than those we observed during the current study. It is possible that this is due to the previous study including a longer treatment period (12 weeks), as well as direct interaction with a physical therapist, which the intervention in our study did not include. It is also possible that these differences in outcomes are related to the specific diagnoses (e.g., impingement syndrome versus adhesive capsulitis) included in each study sample, which was not reported in the study by Janela et al and is not included in our dataset. Additional research is needed to examine how different approaches to mobile application integration (e.g., DETA alone, DETA-physical therapy hybrid) effect patient outcomes.

Interestingly, our results indicate that patients in our cohort with knee pain experienced superior changes in physical function compared to those with shoulder pain. While it is possible that a DETA is more effective in improving function among patients with knee versus shoulder pain, there are a number of other factors that may have contributed to these results. For example, our study did not include specific diagnoses, only the body region being treated. It is possible that the knee pain portion of our cohort included a higher proportion of patients with diagnoses amenable to exercise interventions (e.g., patellofemoral pain) and that patients in our cohort with shoulder pain included a higher proportion of those with diagnoses less amendable to exercise (e.g., adhesive capsulitis). It is also possible that this difference is based on our methods for measuring physical function. Patients with knee pain completed the PROMIS-Physical Function and patients with shoulder pain completed the PROMIS-Upper Extremity Function. While these two measures have been shown to result in very similar T-scores,^15^it is possible that the PROMIS-Physical Function had greater responsiveness in this cohort.

This study includes some of the first indications as to what patient groups may respond best to mobile application interventions for musculoskeletal pain. Notably, we saw the greatest improvements in pain interference and function among those with acute symptoms, those with higher education and those who report higher levels of exertion with exercise. In combination, these findings begin to illustrate the patient profile most suited for mobile application interventions, such as the DETA intervention provided in this study. This patient profile may best be described as the “low risk patient” and could be useful in identifying patients who achieve clinically meaningful outcomes utilizing more independent approaches to rehabilitation, versus in-clinic care. However, more research is needed to continue to develop these types of patient profiles, which are needed in to tailor care-delivery mechanisms (i.e., mobile applications, video visits, in-person care) to the needs of individual patients. This is a particularly important area of research as the field of telerehabilitation continues to expand, including the use of remote therapeutic monitoring which includes the delivery of home exercises through a digital treatment platform and is now recognized as a reimbursable service under Medicare and many commercial insurances.^16^

This study has limitations that should be considered when interpreting its results. First, this study does not include a control arm which limits our ability to examine whether the changes reported by patients in this study were spontaneous or due to the provided interventions.. In addition, we did not have data that could be used to assess patient engagement with the mobile application used in this study which further limits our ability to examine whether utilization of the digital self-management program influenced the outcomes reported by patients in this study. Also, this study is comprised of patients who elected to use the mobile application as an alternative to in-person physical therapy when outpatient visits were restricted due to COVID surges. We do not have information on patients who chose not to utilize the mobile application during this time period. It is possible they differ from our study sample and that the results of our study sample, as a result, are not generalizable to all patients with knee and shoulder pain. Next, our analysis was not restricted to those with specific diagnoses nor did we include this information in our analysis. It is possible that certain diagnoses respond differently to a DETA. Finally, while the intention of the DETA was for it to be used in place of physical therapy, the retrospective nature of this study did not allow us to restrict patients from seeking out physical therapy services outside our institution.

## Data Availability

All data produced in the present study are available upon reasonable request to the authors

